# A Standardised Protocol for Neuro-endoscopic Lavage for Post-haemorrhagic Ventricular Dilatation: A Delphi Consensus Approach

**DOI:** 10.1101/2022.03.20.22272667

**Authors:** DOLPHIN-UK (Developmental Optimisation using Lavage in Preterm Intraventricular Haemorrhage In Neonates) Collaborators, DOLPHIN-UK Collaborators, Kristian Aquilina, Conor Mallucci, Aswin Chari, Saniya Mediratta, Gnanamurthy Sivakumar, Greg James, Ibrahim Jalloh, John Kitchen, Matthew A Kirkman, Patricia de Lacy, Paul Leach, Shailendra Ashok Magdum, William Dawes, William B Lo

**Affiliations:** Department of Neurosurgery, Great Ormond Street Hospital for Children NHS Foundation Trust, Great Ormond Street, London, UK; Department of Paediatric Neurosurgery, Alder Hey Children’s NHS Foundation Trust, Liverpool, UK; Department of Neurosurgery, Great Ormond Street Hospital, London, UK; Department of Paediatric Neurosurgery, Leeds General Infirmary, Great George Street, Leeds, UK; Great Ormond Street Hospital for Children, Great Ormond Street, London, UK; Division of Neurosurgery, Department of Clinical Neurosciences, University of Cambridge, Cambridge, UK; Department of Neurosurgery, Royal Manchester Children’s Hospital, Manchester, UK; Department of Neurosurgery, Queen’s Medical Centre, Nottingham, UK; Department of Paediatric Neurosurgery, Sheffield Children’s NHS Foundation Trust, Sheffield, UK; Department of Neurosurgery, University Hospital of Wales, Cardiff, UK; Department of Neurosurgery, Oxford University Hospitals NHS Foundation Trust, Oxford, UK; Department of Neurosurgery, Alder Hey Children’s Hospital, Liverpool, UK; Department of Neurosurgery, Birmingham Women’s and Children’s Hospital NHS Foundation Trust, Birmingham, UK

**Author notes:** Corresponding Author: saniya Mediratta.

**Keywords:** Neuro-endoscopic lavage, post-haemorrhagic ventricular dilatation, post-haemorrhagic hydrocephalus, Delphi consensus, neurodevelopmental outcome

## Abstract

**Purpose:** Neuro-endoscopic lavage (NEL) has shown promise as an emerging procedure for intraventricular haemorrhage (IVH) and post-haemorrhagic ventricular dilatation (PHVD). However, there is considerable variation with regards to the indications, objectives, and surgical technique in NEL. There is currently no randomised trial evidence that supports the use of NEL in the context of PHVD. This study aims to form a consensus on technical variations in the indications and procedural steps of NEL.

**Methods:** A mixed methods modified Delphi consensus process was conducted between consultant paediatric neurosurgeons across the United Kingdom. Stages involved literature review, survey, focused online consultation and iterative revisions until > 80% consensus was achieved.

**Results:** Twelve consultant paediatric neurosurgeons from 10 centres participated. A standardised protocol including indications, a 3-phase operative workflow (pre-ventricular, intraventricular, post-ventricular) and post-operative care was agreed upon by 100% of participants. Case- and surgeon-specific variation was considered and included through delineation of mandatory, optional, and not recommended steps.

**Conclusion:** Expert consensus on a standardised protocol for NEL was achieved, delineating the surgical workflow into three phases: pre-ventricular, intraventricular, and post-ventricular, each consisting of mandatory, optional, and not recommended steps. The work provides a platform for future trials, training, and implementation of NEL.

## Introduction

Neuro-endoscopic lavage (NEL) is a relatively novel adjunct to the treatment for post-haemorrhagic ventricular dilatation (PHVD) in preterm neonates that aims to improve cognition and neurodevelopmental outcomes by early removal of blood and its toxic breakdown products that may affect normal brain development [1]. It was formally added as a new recommendation in Part 2 of the 2014 Congress of Neurological Surgeons Paediatric Hydrocephalus Guidelines as a feasible and safe option for the removal of intraventricular clots [2].

Retrospective studies have suggested that NEL may be effective at reducing shunt dependence and allowing good motor and cognitive outcomes [3–7]. However, there is considerable variation in NEL practice and currently there is no randomised trial evidence supporting the use of NEL [8].

In this study, a modified Delphi consensus was conducted to standardise the inclusion criteria, operative workflow, and early post-operative care of NEL, with the aim of ensuring the procedure is delivered consistently and uniformly across centres. Similar modified Delphi methodologies have been effective in developing workflows for contemporary surgical procedures [9–15]. The work provides a platform for future trials, training, and implementation of NEL [10, 14, 15].

## Methods

To achieve a comprehensive protocol for NEL, expert opinion was obtained through an iterative process (Figure 1) to produce an operative workflow for 3 phases of NEL: pre-ventricular, intraventricular, and post-ventricular, as well as the indications for the procedure and post-operative care.

**Fig. 1.**
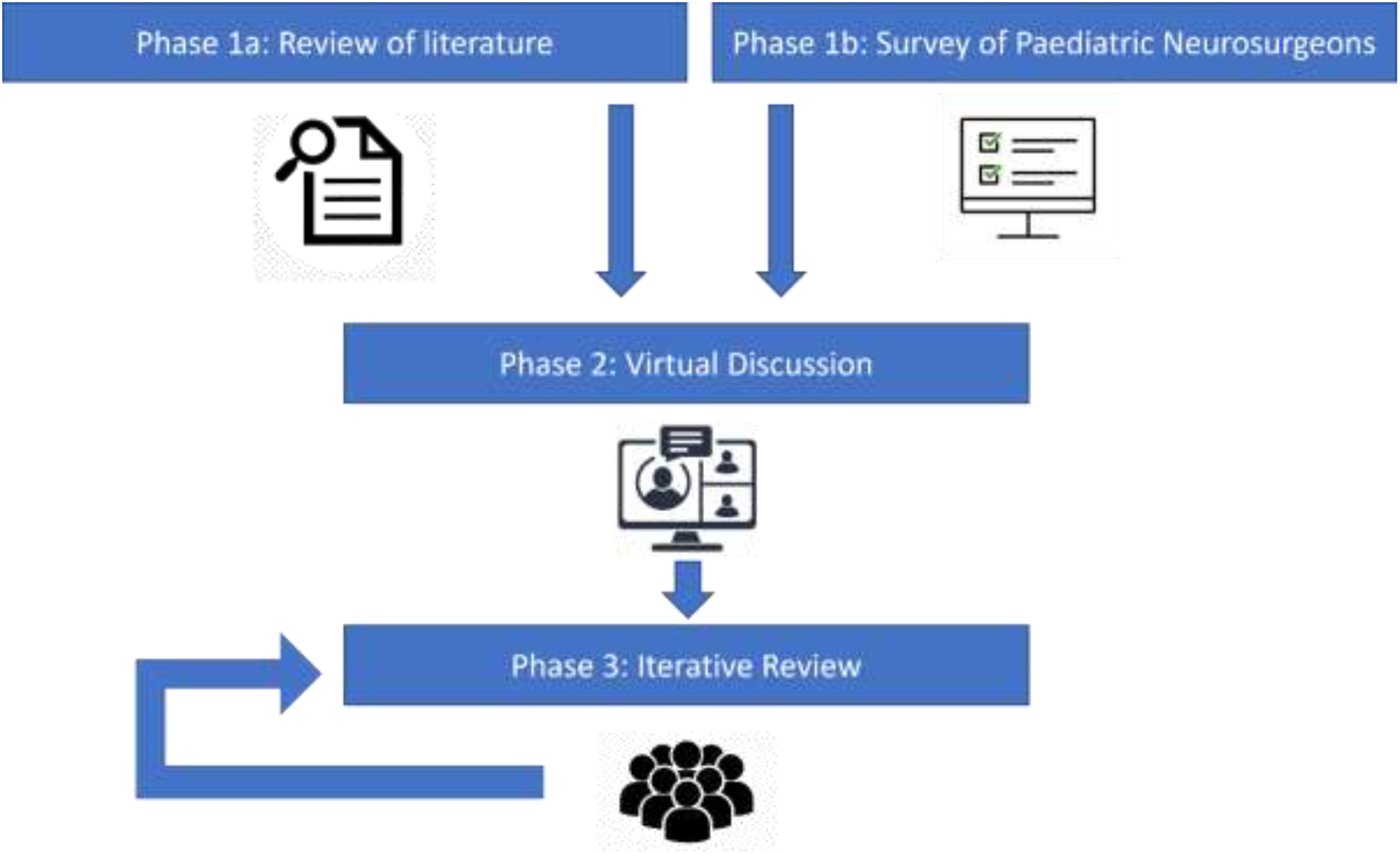
Schematic of mixed-methods Delphi consensus process.

### Consensus Phase 1

A two-step approach was used to create a preliminary operative workflow for NEL.

#### Phase 1a: Literature Review

To create a preliminary framework for the operative workflow, a literature review was conducted using PubMed. The search criteria included 5 keywords: “neuro-endoscopic lavage”, “NEL”, “ventricular lavage”, “post-haemorrhagic ventricular dilatation”, and “post-haemorrhagic hydrocephalus”. An initial workflow was created using the articles found, and areas of clarification, discussion and opinion were highlighted based on technical variation found within these resources.

#### Phase 1b: Consensus Sampling

Alongside the literature review process, fully trained consultant paediatric neurosurgeons with an interest in NEL were selected to be surveyed on their opinions or experience with the procedure. All neurosurgeons had completed a clinical fellowship in paediatric neurosurgery during their training. The panel of neurosurgeons were selected to ensure members had (i) experience in different tertiary paediatric neurosurgery institutions across the United Kingdom and (ii) experience managing and operating on infants with PHVD. A 12-question web-based survey was designed, including questions on inclusion and exclusion criteria, operative workflow, and post-operative care (*Supplementary Figure 1)*.

### Consensus Phase 2

The literature review and anonymised survey results of Phase 1 were presented back to the group of paediatric neurosurgeons through an online, video-teleconferencing forum. Each member was invited to pose questions for discussion and factors for consideration. All suggestions were documented for incorporation to the protocol if they were (i) relevant to the aims of the NEL protocol and (ii) received agreement from > 50% of the consensus group present at the meeting.

### Consensus Phase 3

Using the information from Phase 1 and 2, a suggested protocol was formed outlining inclusion and exclusion criteria, operative workflow, and post-operative care for NEL. Each operative step was delineated as mandatory, optional, and not recommended based on results of Phases 1 and 2. According to the Delphi technique, circulation and iterative revision of the protocol was repeated until all experts were in agreement that the protocol was comprehensive and accurate. Factors that did not achieve 80% consensus were included as suggestions and highlighted as areas for further research.

## Results

### Phase 1a: Literature Review

Eight original articles were identified with reports and experiences of using endoscopic lavage for removal of blood products in preterm haemorrhage. Seven of these studies were retrospective cohort studies (Table 1) and one was a descriptive study highlighting technical experience in NEL. All surgical descriptions used similar protocols for endoscopic lavage with technical variations that were used to construct the initial operative workflow and highlight points of discussion and discrepancies in protocols for Phases 1b and 2.

**Table 1:**
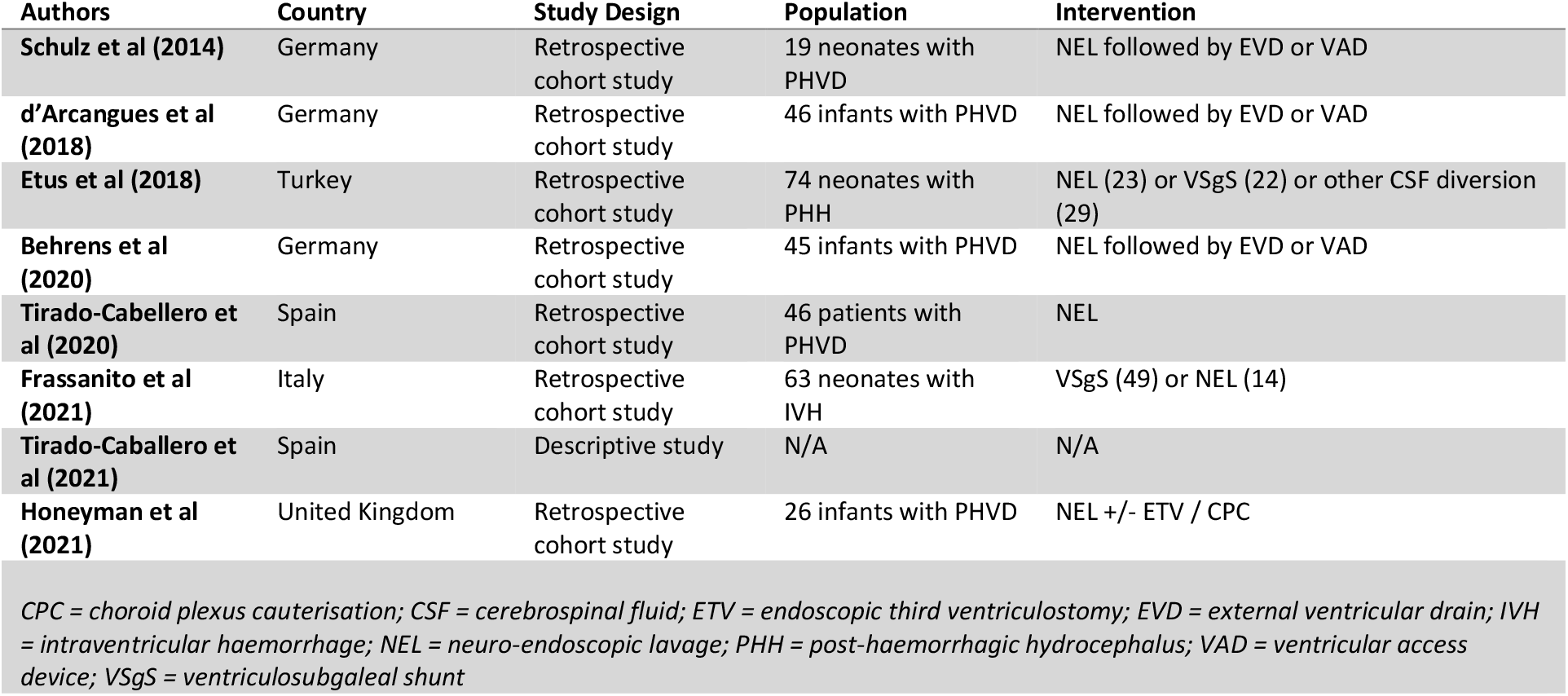
Summary of articles identified on NEL [1, 3–8].

| Authors | Country | Study Design | Population | Intervention |
| --- | --- | --- | --- | --- |
| Schulz et al (2014) | Germany | Retrospective cohort study | 19 neonates with PHVD | NEL followed by EVD or VAD |
| d'Arcangues et al (2018) | Germany | Retrospective cohort study | 46 infants with PHVD | NEL followed by EVD or VAD |
| Etus et al (2018) | Turkey | Retrospective cohort study | 74 neonates with PHH | NEL (23) or VSgS (22) or other CSF diversion (29) |
| Behrens et al (2020) | Germany | Retrospective cohort study | 45 infants with PHVD | NEL followed by EVD or VAD |
| Tirado-Cabellero et al (2020) | Spain | Retrospective cohort study | 46 patients with PHVD | NEL |
| Frassanito et al (2021) | Italy | Retrospective cohort study | 63 neonates with IVH | VSgS (49) or NEL (14) |
| Tirado-Caballero et al (2021) | Spain | Descriptive study | N/A | N/A |
| Honeyman et al (2021) | United Kingdom | Retrospective cohort study | 26 infants with PHVD | NEL +/- ETV / CPC |

CPC = choroid plexus cauterisation; CSF = cerebrospinal fluid; ETV = endoscopic third ventriculostomy; EVD = external ventricular drain; IVH = intraventricular haemorrhage; NEL = neuro-endoscopic lavage; PHH = post-haemorrhagic hydrocephalus; VAD = ventricular access device; VSgS = ventriculosubgaleal shunt

### Phase 1b: Consensus Sampling

12 paediatric neurosurgeons (*Supplementary Figure 2*) were selected to participate, with a 100% response rate in Phase 1b. 16.7% (n=2) of respondents believed NEL should be performed on all IVH, whereas 25% (n=4) responded it should be performed on Grade II and above only, 41.7% (n=5) responded Grade III IVH and above, and 16.7% (n=2) responded Grade IV only. The majority (58.3%, n=7) reported that NEL should be performed on development of PHVD, defined as an increase in the ventricular index beyond the 97^th^ centile + 4mm, whilst 25% (n=4) defined this point as when the VI crosses the 97^th^ centile. 83.3% (n=10) of respondents agreed that NEL should be delayed until the infant is systemically stable.

The majority of respondents (83.3%, n=10) reported that either a flexible or rigid endoscope could be used. With regards to which ventricle should be entered first, 66.7% (n=8) felt the ventricle with the higher clot burden should be entered first, 25% (n=3) reported that it does not matter and 8.3% (n=1) felt the ventricle with the lower clot burden should be entered. There was no preference between the left or right ventricle. 100% of respondents agreed that the frontal horn of the ventricle should be cannulated.

With regards to lavage, 58.3% (n=7) would use Ringer’s lactate as the irrigant, 25% (n=4) favoured artificial CSF and the remainder of respondents (16.7%, n=2) did not believe the type of irrigant made a difference. No respondents selected Plasmalyte as the optimal fluid. 41.7% (n=5) did not believe a pre-determined quantity of irrigant was required. The options of 1 litre or 2 litres of irrigant were selected by 1 respondent each. The remaining respondents selected “other” and through free text suggested that enough should be used to clear the clot (8.3%, n=1) or until the effluent was clear (8.3%, n=1), raising considerations such as intra-operative stability (25%, n=3). This corresponded to the majority of respondents reporting that the endpoint of lavage should be once the effluent fluid is clear or once all the clots have been removed (58.3% and 16.7% respectively).

Additional steps recommended by respondents to be included as part of the procedure where possible are shown in Figure 2a, along with responses about the use of temporising devices following NEL (Figure 2b). 91.7% (n=11) would give prophylactic antibiotics at induction, 41.7% (n=5) would also give prophylactic antibiotics for 24 to 48 hours post-operatively. No respondents would administer intrathecal antibiotics at the end of the procedure.

**Fig. 2.**
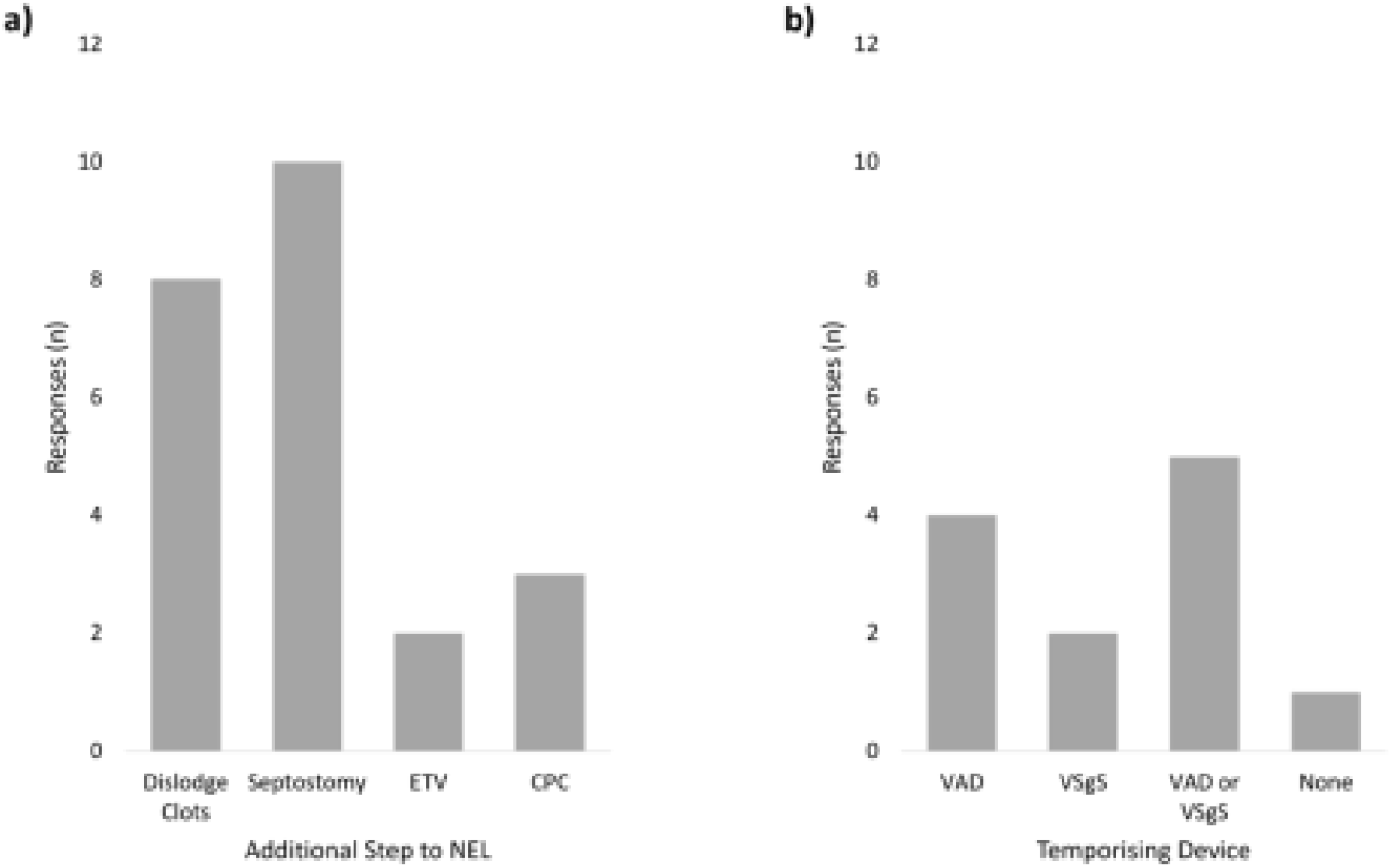
Survey responses to additional steps to be included as part of neuro-endoscopic lavage (NEL) (A) and procedure to be performed followed NEL (B). CPC = choroid plexus cauterisation; ETV = external ventricular drain; VAD = ventricular access device; VSgS = ventriculosubgaleal shunt.

#### Consensus Phase 2

7 neurosurgeons participated in the group discussion where the results of Phase 1 were presented. Questions in which there were differences in opinion were discussed and explained, such as, the inclusion criteria, the use of temporising device following NEL and the endpoint for lavage. The results of Phase 1 along with descriptive results of Phase 2 were circulated to the remaining participants to ensure they had the opportunity to raise any additional points for discussion or suggestions. During Phase 2, it was agreed that the structure of the final protocol would be categorised into three operative phases (pre-ventricular, intraventricular, and post-ventricular) and the steps within each phase would be delineated as mandatory, optional, or not recommended.

#### Consensus Phase 3

During Phase 3, multiple iterations were performed, and 100% consensus was achieved on the final protocol as described below and summarised in Figure 3.

**Fig. 3.**
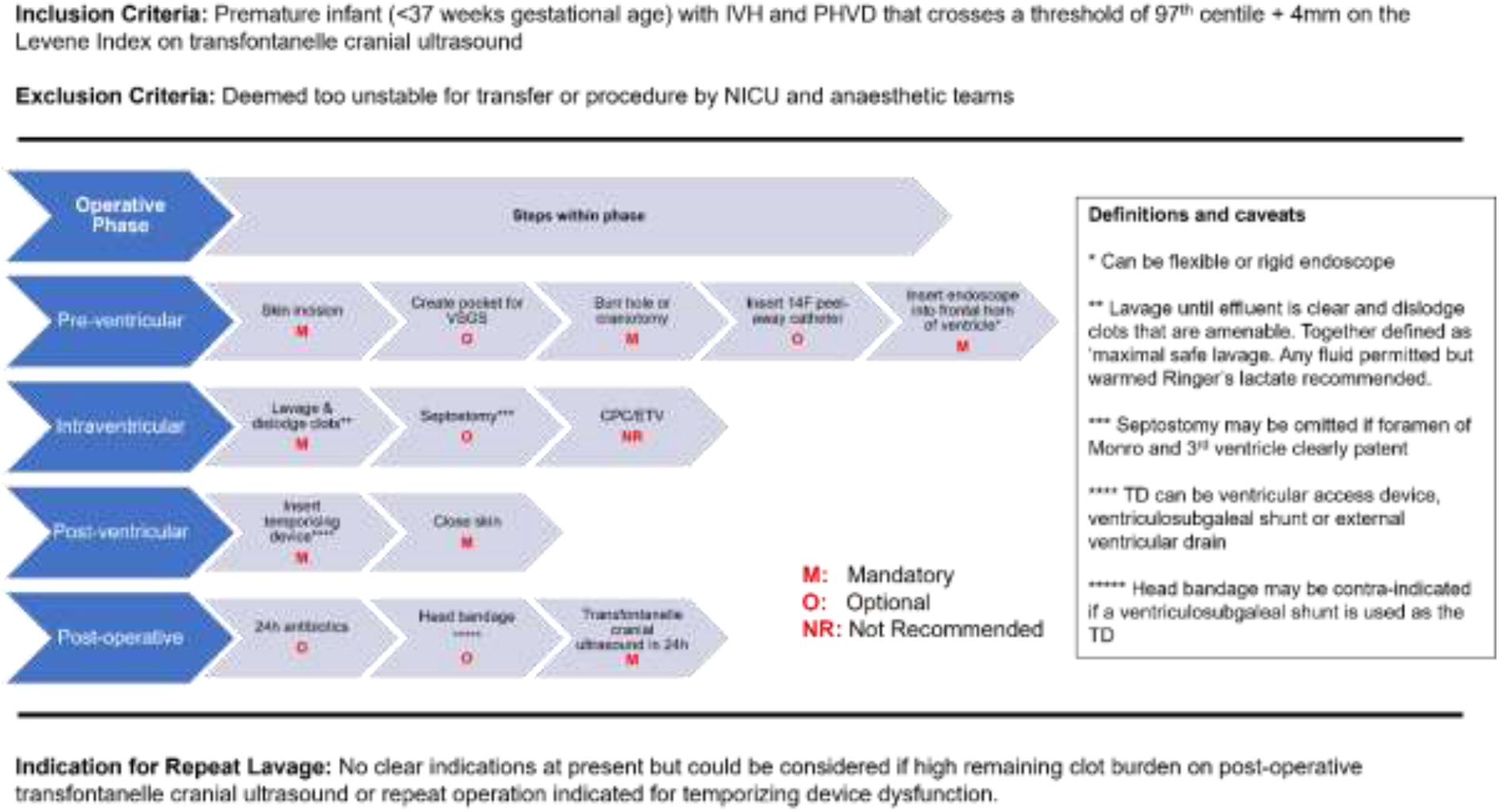
Consensus-derived protocol for neuro-endoscopic lavage in post-haemorrhagic ventricular dilatation.

### Inclusion and Exclusion Criteria

The inclusion criteria for NEL were premature infants, defined as gestational age less than 37 weeks, with IVH and PHVD that crosses a threshold of 97th centile + 4mm on the Levene Index of transfontanelle cranial ultrasound[16]. NEL was not recommended in cases where the infant was deemed too unstable for either (i) transfer or (ii) the procedure, by neonatal ICU and anaesthetic teams.

### Pre-Ventricular Phase

This phase was defined from the point of skin incision to initial entry into the ventricle with an endoscope, which was defined as being either a fixed or a rigid endoscope depending on surgeon preference and endoscope availability. There were 5 steps in the pre-ventricular phase of which 3 were mandatory, and 2 optional.

### Intraventricular Phase

The intraventricular phase included one mandatory step which was the lavage and dislodging of clots. 100% agreement was achieved on the endpoint of ‘maximal safe lavage’, defined as lavage until effluent is clear and all of the clots that are amenable to removal have been dislodged. Septostomy was included as an optional step depending on the case (e.g. patency of the foramen of Monro and third ventricle and visibility of septum pellucidum), and ETV and CPC were not recommended.

### Post-ventricular Phase

The post-ventricular phase was composed of 2 mandatory steps: insertion of a temporising device and skin closure. The specific type of temporising device was not defined, in acknowledgement of the heterogeneity of device choice based on case and surgeon or institutional preference [17].

### Post-operative Management

The consensus group recognised that post-operative care varied significantly based on surgeon preference, and therefore 2 optional steps were included in the protocol of administration of antibiotics within 24 hours and placement of a head bandage post-operatively (especially as this may occlude the subgaleal pocket if a VSgS was chosen).

No distinct indications were identified for repeat NEL; however, it was agreed that this may be indicated in specific circumstances such as, (i) repeat lavage if repeat operation was already being performed for temporising device dysfunction or (ii) if a high residual clot burden was visualised on post-operative transfontanelle cranial ultrasound.

## Discussion

A standardised protocol for NEL was generated using a modified mixed-methods Delphi technique with national expert consensus.

Whilst the principles of ventricular lavage in PHVD have remained consistent over time, there has been variation in the techniques proposed to deliver ventricular lavage [2, 18–20]. Advantages have been described which favour NEL, for example, allowing direct visualisation of the progress of haematoma removal and therefore enabling targeted irrigation and clot removal under direct vision[1]. Another advantage is that this procedure is often delivered during the time of insertion of a temporising device within the same operative session, avoiding additional risks associated with repeated procedures and sessions under general anaesthetic [19]. With neuro-endoscopic lavage emerging as a feasible and safe technique, technical variation between operating paediatric neurosurgeons exists [1–8, 19, 20]. Whilst this made the consensus process challenging, the discussion and multiple iterations that arose from the staged consensus process during this study were valuable and relevant. The incorporation of optional steps within each phase of protocol acknowledges important variations in practice between individual surgeons and institutions as well as importantly providing scope for adaptation of the protocol to specific cases.

Components of the protocol that were not mandated or specified, such as type of temporising device, reflect areas where there is limited evidence to suggest that these factors have any influence on outcome, highlighting areas of practice that would benefit from future research should NEL be validated and implemented as routine management for PHVD. Interestingly, despite variation in the published practice of NEL [1, 3–5, 7, 8], all participating paediatric neurosurgeons in this consensus ultimately agreed that NEL should be performed as an adjunct and not replacement to the procedure of temporizing device insertion, particularly when NEL is being performed at the time of development of PHVD (when the ventricular index crosses the 97^th^ centile + 4mm).

### Applications

The benefits of generating a standardised workflow and protocol for novel procedures such as NEL are well described in the literature in applications within neurosurgery and across surgical specialties [9, 11–13]. Firstly, the protocol developed will provide standardisation for the development and implementation of a national randomised-controlled trial to investigate the efficacy of NEL in PHVD. In this context, it can also play a role in quality control and assessment of procedural variability across the neurosurgical units participating in the trial. Secondly, if this technique is to be implemented into routine best-practice management for PHVD, this structured, operative workflow will be a valuable tool in education and training for the procedure at the point of implementation [12, 13, 21, 22]. Finally, the development of a structured, logical surgical workflow will form a useful tool when explaining the procedure to parents of children with PHVD. The inclusion of mandatory steps and optional steps allows clear communication of the case-specific aspects of the procedure within the process of information sharing and consent with parents and carers.

Importantly, the process undertaken in this study reflects critical stages of the IDEAL (Idea, Development, Exploration, Assessment and Long-term study) Framework, a well-described framework for surgical therapy innovation [23]. Refining technical details of a surgical procedure through experiences of small case series was effectively conducted herewith and is described as the Development Stage 2a of the IDEAL framework [24]. The Exploration Stage outlines the process of reaching a collective understanding of the procedure between operators from different centres, to address obstacles to a definitive comparative trial, including reaching agreement about standard technique and accepted variants. This study highlights this crucial step that should be a requirement in the development of neurosurgical innovation [25, 26]. Accordingly, the NEL protocol produced through this consensus study will serve as valuable tool in the design of a collaborative randomised clinical trial to determine the efficacy of NEL, in line with Stage 3 of the IDEAL Framework.

### Limitations

It is important to highlight the limitations due to the scope of this study. Firstly, whilst the Delphi method was applied to a national, multi-centre panel, international variations in practice were not accounted for or incorporated in phases 1b to 3 and would add value in the future. Published international experience was accessible and utilised during the literature review phase [4, 7, 8]. The Delphi method is a well-described and effective technique for obtaining consensus between groups of experts, however, the protocol generated from this process remains at the level of expert opinion based on experience of the participants [11, 14, 15, 27]. Further prospective, objective studies into each individual recommendation once the practice of NEL is more established remains essential. Furthermore, variations in pre-operative protocols were not covered in the scope of this study, such as, surgical positioning or administration of thrombolytic agents [8].

## Conclusion

A standardised protocol for NEL was generated using a modified mixed-methods Delphi technique with national expert consensus, structuring the procedure into three phases (pre-ventricular, intra-ventricular and post-ventricular) consisting of mandatory, optional, and not recommended steps. The operative workflow generated will provide a template for future research, education, and implementation of NEL in the context of PHVD. Furthermore, the consensus process highlighted technical variations which would benefit from further research, such as specification of the fluid used for lavage.

## Data Availability

The data that support the findings of this study are available from the corresponding author [SM] upon reasonable request.

## Abbreviations

CPC: Choroid plexus cauterisation
CSF: Cerebrospinal fluid
EVD: External ventricular drain
ETV: Endoscopic third ventriculostomy
IVH: Intraventricular haemorrhage
NEL: Neuro-endoscopic lavage
PHH: Post-haemorrhagic hydrocephalus
PHVD: Post-haemorrhagic ventricular dilatation
VAD: Ventricular access device
VSgS: Ventriculosubgaleal shunt

---

**Supplementary Figure 1:**
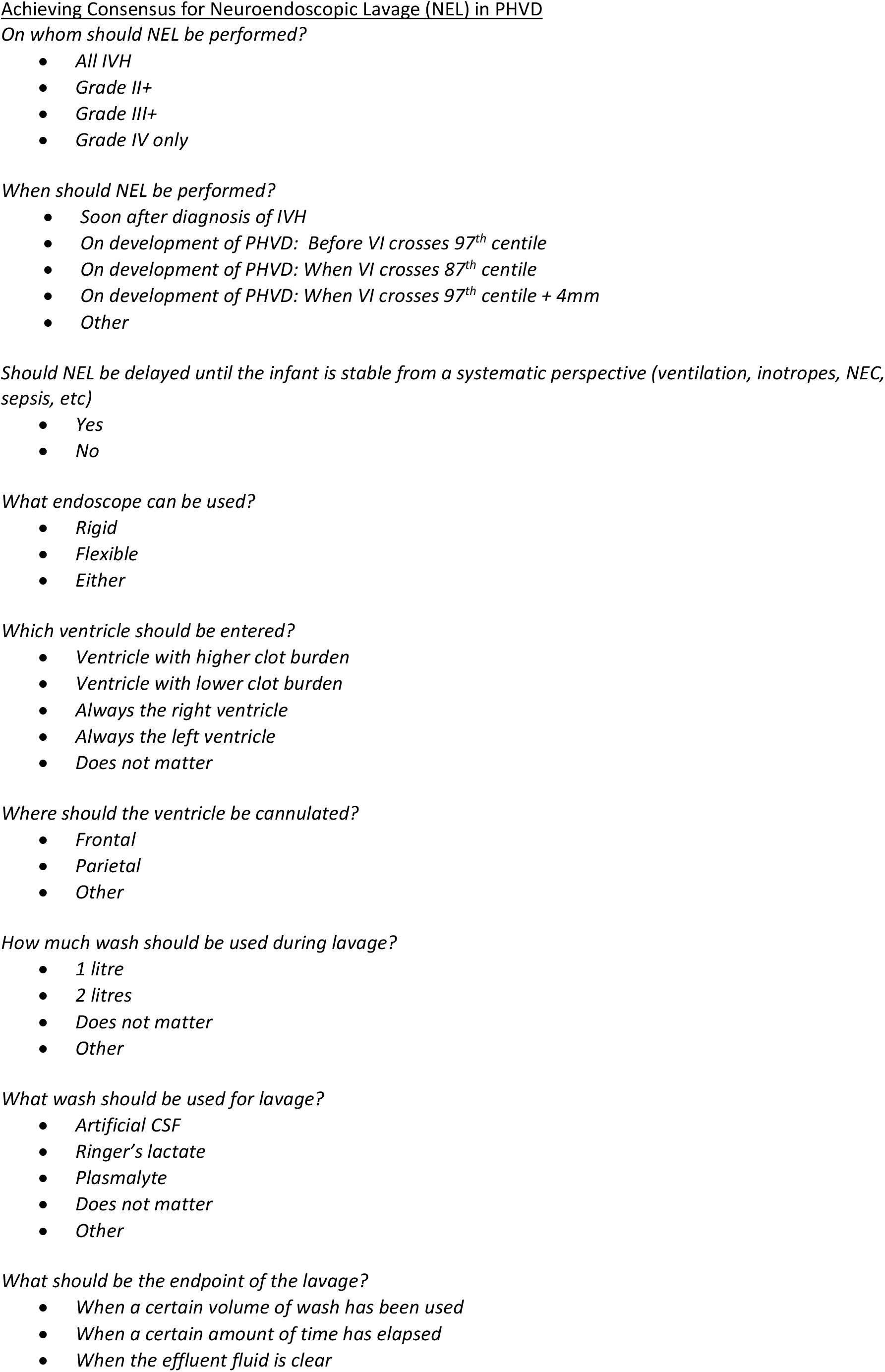
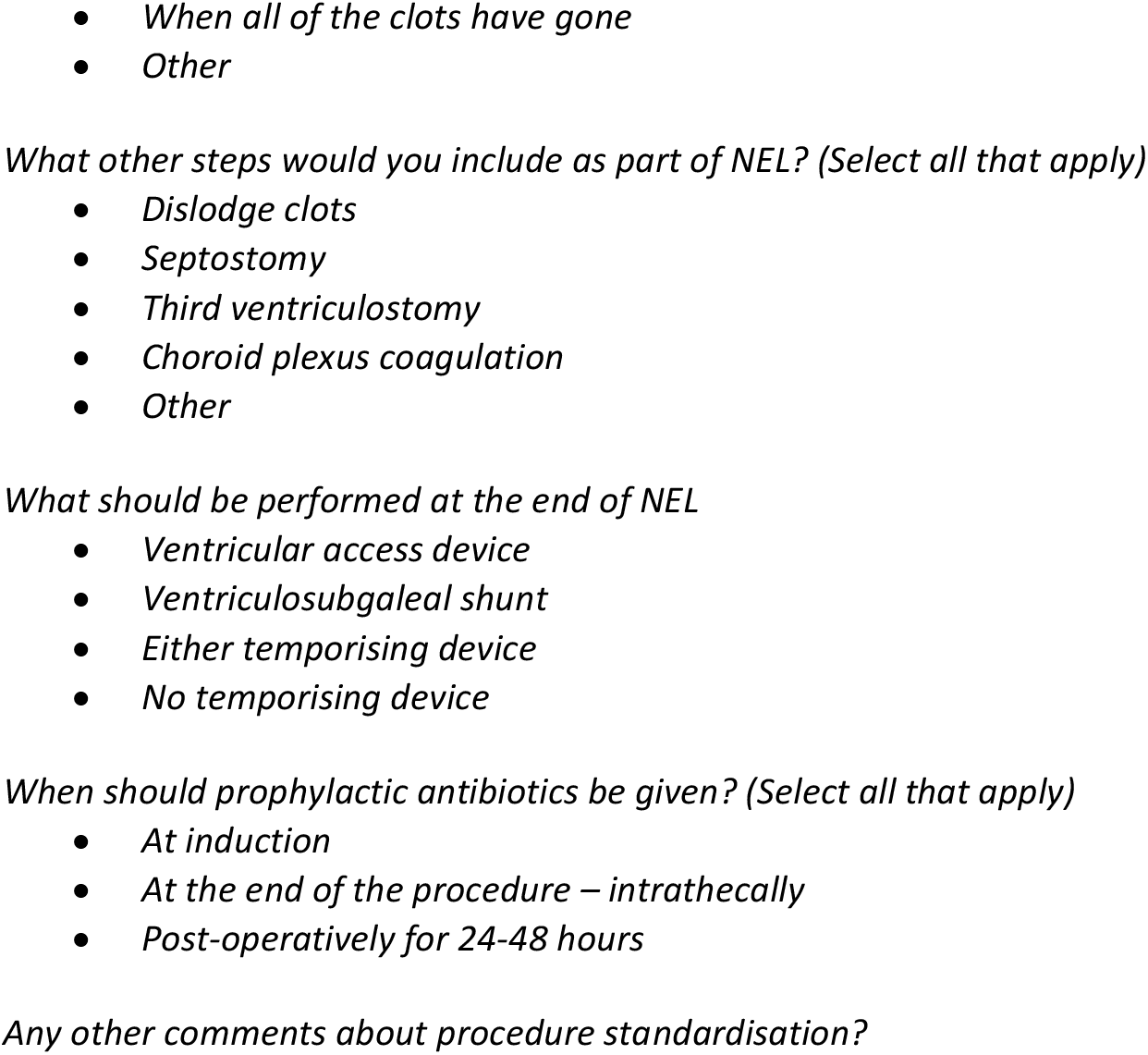
Phase 1b Survey.

**Supplementary Figure 2:**
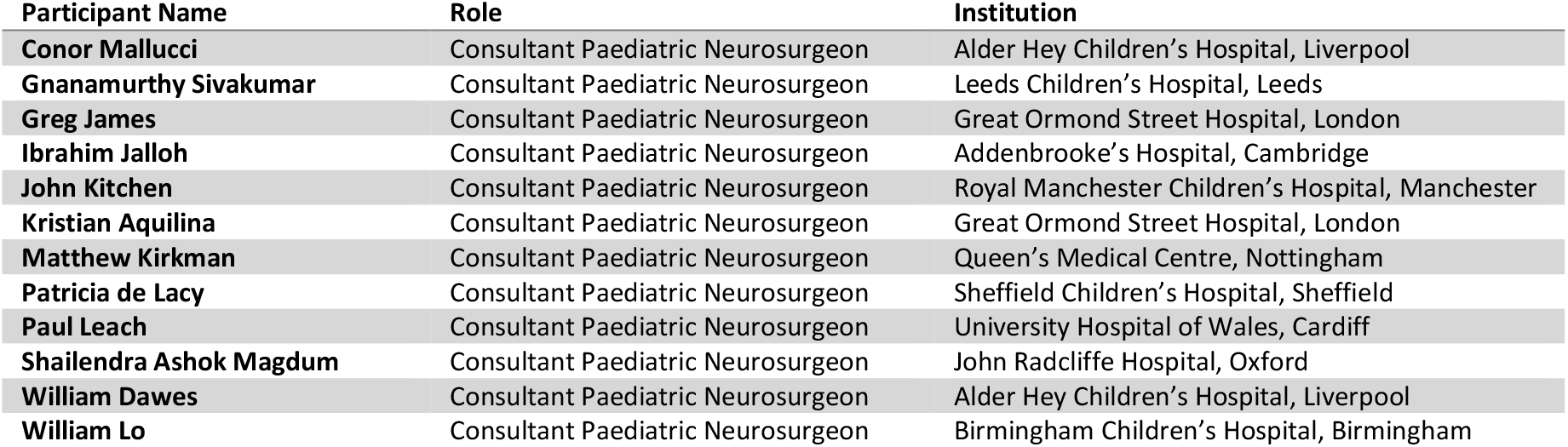
Consensus Participants.

## Author Contributions

Conceptualization: KA, AC, SM; Methodology: AC, SM, KA; Data Collection: AC, SM, KA, CM, GS, GJ, IJ, JK, MAK, PL, SAM, WD, WBL; Formal analysis and Investigation: SM, AC, KA; Writing - original draft preparation: SM, Writing - review and editing: AC, KA, SM; Manuscript Review: CM, GS, GJ, IJ, JK, MAK, PL, SAM, WD, WBL; Supervision: AC, KA

